# Menstruation Hygiene Management among Secondary School Students of Chitwan, Nepal- Pre-Post Study Design

**DOI:** 10.1101/2022.12.05.22283126

**Authors:** Gayatri Khanal, Niki Shrestha, Kishor Adhikari, Usha Ghimire

## Abstract

**Introduction:** Despite of significant progress towards reproductive health, still in many societies menstruation is treating as negative, shameful or dirty consequences which may increases the incidence of reproductive tract infection leading to significant negative impact to a women’s health. To manage menstruation hygienically and with dignity, it is essential that women and girls have sufficient knowledge on menstruation hygiene management to manage it. Thus, this study aim to identify the knowledge and practice of menstruation hygiene and effectiveness of a health educational intervention on MHM.

**Methods:** A school-based cross-sectional study was conducted between August 2021 to April 2022 among 400 secondary school girls studying in grades 9 and 10 in Chitwan District, Nepal. A total of 20 schools out of 250 schools were selected through lottery method. A further 400 girls were selected from selected schools by using systematic random method. Logistic regression analysis and Wilcoxon rank test were employed to identify predictors and evaluate effectiveness of health educational intervention on Menstrual Hygiene Management

**Results:** Overall, 57.7% of the girls had unsatisfactory level of knowledge. Around two third (61.4%) missed schools days. Almost 99.5% had experienced any form of cultural restrictions. After health education intervention, level of knowledge about menstruation hygiene had significantly improved (z= 17.129, p=<0.001) in satisfactory level of knowledge compared to baseline (42.3% vs 92.5%). During multivariate analysis, studied in public schools (AoR= 1.7, p= 0.026), No or one close female friend (AoR= 2.2, p= 0.011), other than Brahmin/Chhetri caste (AoR= 1.4, p= 0.05), lived in joint family (AoR= 1.6, p= 0.048) were significantly associated with unsatisfactory level of knowledge on Menstruation hygiene management.

**Conclusion:** Girls had unsatisfactory level of knowledge. School absenteeism and cultural restriction is very high. Ethnicity status, Types of family, Number of close female friends, types of schools and mothers’ education were associated with MHM.. Educational program conducted brings significant change in level of knowledge. Hence the education intervention demonstrate the suitability and feasibility of implementing a health educational intervention on MHM.

## Introduction

Menstruation is a natural process and a key sign of reproductive health, yet, in many societies it is treated as something negative, shameful or dirty. The continued silence around menstruation combined with limited access to information at home and in schools results in millions of women and girls having very little knowledge about what is happening to their bodies when they menstruate and how to deal with it.^1, 2^ A study revealed that one out of three girls in South Asia knew nothing about menstruation prior to getting it. ^1^

Globally, approximately 52% of the female population (26% of the total population) is within reproductive age. Most of these women and girls will menstruate each month for between two and seven days. To manage menstruation hygienically and with dignity, it is essential that women and girls have sufficient knowledge on menstruation hygiene management (MHM) to manage it. ^3^

Every year approximately, 10% of women worldwide are exposed to genital infections including urinary tract infections and bacterial vaginosis, and 75% of women have a history of a genital infection. Specifically, common risk factors for vaginal infections include pregnancy and poor hygiene (both perineal and menstrual hygiene). ^4^ The knowledge and practice of good menstrual hygiene reduces the incidence of reproductive tract infection (RTI). The consequences of RTI’s are severe and may result in significant negative impact to a women’s health including chronic pelvic pain, dysmenorrhea (painful periods) and in severe cases, infertility. Reproductive tract infections, which have become a silent epidemic that devastates women’s lives is closely related to poor menstrual hygiene. ^5, 6^

Every day, approximately 2, 90,000 women and girls in Nepal experience menstruation. With 82% of Nepali women living in rural Nepal, use of archaic, unhygienic, unhealthy and possibly dangerous menstrual hygiene management methods push Nepali women deeper into the crevice of marginalization and reproductive health morbidity. ^7^ The practical challenges of menstrual hygiene are made even more difficult by the socio-cultural factors in Nepal. ^8^ Many girls and women face challenges with managing their periods safely because of persisting taboos. For example, the custom of “Chhaupadi” is still prevalent in Nepal – a deplorable custom which relegates women to live in a cow shed and experience grossly unhygienic conditions while menstruating.

Though, Nepal government has done significant progress towards reproductive health, adolescent girls still face many sexual and reproductive health problems and challenges, amidst gender discrimination and disparity in Nepali norms.

Young adolescent girls constitute a particularly vulnerable group. Adolescent girls often lack appropriate information about their reproductive health and proper menstrual management which has a direct impact on daily living leading to school absenteeism as high as 53% in Nepal. This implies an average annual absence of at least six weeks from school directly affecting girl’s academic career. Unsatisfactory academic progress linked with the societal belief that girls who have started menstruating are eligible for marriage, contributes to high dropout rate and early marriage which itself is a factors for preventing optimum reproductive health, birthing practices, birth spacing and healthy children. ^7, 8^ To reduce such problems small effort like awareness to adolescent girls can play crucial role. ^7, 8, 9, 10^

It has been reported that females who were aware of menstruation hygiene and stayed longer in school is associated with reduced maternal death; improved population health; increased contraceptive uptake; decreased fertility rate, improved child health and decreased infections. ^10^ Interventions that increases awareness of menstruation hygiene may clearly have important secondary health outcomes and wider economic benefit. Thus, this study was conducted to identify the knowledge and practice of menstruation hygiene and effectiveness of a health educational intervention on MHM.

## Methods

### Study design, study settings and study participants

School based cross-sectional study was organized from May 2021 to April 2022 among secondary schools girls in Chitwan District, Nepal. Seclusion and exclusion practices were widespread, particularly among Brahmin, Chhetri and Newar cast and these group of cast predominantly reside in the Chitwan. ^11^ Secondly, this district was selected to represent different terrains as well as rural –urban areas. ^12^ In Chitwan district, there were 250 secondary schools among them 122 schools were public and 128 were private. There were 13652 girls enrolled in the 9^th^ and 10^th^ grade for the academic year 2020-2021 out of which 9344 were in public schools and 4308 were in private schools. ^13^ The girls who had attained menarche were included while those who were absent on the day of data collection were excluded from the study.

### Sample size and sampling procedure

The sample size was determined using population proportion formula given by Cochran with the assumption of 95% confidence, 5% allowable error, and prevalence of knowledge about menstruation hygiene at 39.2%. ^14^ To compensate for the non-response rate, 10% of the determined sample was added up on the calculated sample size and the final sample size was found to be 400.

The sampling procedure was started by selection of the schools. 20 out of 250 were selected from both public and private schools. A lottery method was used to select the secondary schools from the district. For the lottery method, name of all the secondary schools of the district was written in a separate, uniform-sized paper which was folded and put into a box. The papers was thoroughly mixed. One by one, the papers were taken out while the box was thoroughly shaken every time.

Total list of girls (9^th^ and 10^th^ grade) in each secondary schools was obtained from the student registration books of the respective schools. There were 1237 girls, out of which 400 girls were selected by using systematic random sampling procedure. To perform systematic sampling kth items was calculated (1237/400=3.09^th^) and first number was assigned through simple random sampling techniques.

If the selected girl was absent on the day of data collection, the next nearest was chosen as a respondent.

### Data collection Tools and Techniques

The data collection tool was Semi-structure self-administer questionnaire and observation checklist for observation of school environment which was prepared in Nepali language on the basis of a review of the relevant literatures. The tool was developed with the help of similar studies ^15, 16, 17^ and extensive literature review of related documents and help of expertise in the field of MHM.

Before data collection, participants were approached through school authority. Research participants were clearly informed about type of research, objectives, and its benefits in near future. Four last year students of Bachelor of Public Health were recruited as data collectors. They were given one day training and education to familiarize with the study, maintain confidentiality of information and other collection procedures to follow as per program need. Supervision on data collection was done by the researcher team. Initially, self-administered questionnaire was given to perform pretesting then intervention (health education) package was given and finally posttest was done. Pre testing questionnaire covered information on demography, knowledge and practices. Post test questionnaire covered only knowledge related aspects. Interventional (educational) package included the menstrual components like definition, causes, hygienic practices, myths, remedial measures to relive discomfort and pain etc. Data was collection was done from 7^th^ April to 27^th^ April 2021. To ensure the reliability of the tool, pilot testing was done.

Pilot-testing was conducted among 10 % of sample size in 4 high schools (2 public and 2 private) of Chitwan district. The feedbacks from the pilot-testing was incorporated and questionnaire was then finalized. Necessary modifications and improvisation to the questionnaire was done following the test.

## Data analysis

Data entry was done using Epi-data version 4.6. The data was checked for completeness and accuracy before feeding data on Epi-data 3.1. Data analysis was done using IBM Statistical Package for the Social Sciences (SPSS) Version 20.

The statistical analysis carried out was descriptive, bivariate (Wilcoxon sign rank test), multivariate analysis (Logistic regression). Those variables at p value ≤ 0.05 was considered as significant. Before multivariate analysis, bivariate analysis was performed between dependent variable (level of knowledge) and each of independent variables (socio-demographic and other variables), one at a time. The variables which were found significant at bivariate test were channeled into multivariate analysis using a logistic regression model in order to control for confounding factors.

The model for goodness of fit was checked using the Hosmer-Lemeshow test (p >0.05.) and Variation inflation factors (VIF) of all significant independent variables which lies in the range of 1-4. Level of knowledge was calculated out of the 24 knowledge specific questions. (Supplementary file). Each correct response considered as a 1 point and an incorrect response was considered as 0. The highest sum score of knowledge was 24 whereas lowest is 0. The level of knowledge was categorized as per the study conducted by Chetkanta et al., in Dang, Nepal. ^18^Unsatisfactory: 0-12 score; Satisfactory: 13-24.

### Inclusion and exclusion criteria

Only those girls who are currently studying in 9^th^ or 10^th^ grades were included in the study. Those girls who have not experienced menarche, refused informed consent and absent on the day of the data collection were excluded from the study.

### Ethical issues

The Nepal Health Research Council Ethical Review Committee was approached for ethical clearance to carry out this study. Ethical clearance (ERB protocol Registration No 139/2021P) was obtained for this study on 7^th^ April 2021. A written permission for carrying out this study was taken from Education Development and Coordination Unit, Chitwan and the respective secondary schools. Verbal informed consent was obtained from the girls before starting the data collection. The data was kept confidential.

## Results

### Socio-demographic characteristics of the study participants

The participants’ ages were between 12 to 19 years, with a mean age 15 and SD 0f ±1. 87.3% respondents belonged to Hindu religion. Of the total, 50.7% were studying in public schools. Most of the schools (95.8%) were located near the market areas. 82.2% girls had more than one close friend. 12.5% mothers of the participants were illiterate.

### Menstruation related information

More than half (53.2%) girls had never participated in MHM related program. Majority of the girls (85%) had heard about menstruation before menarche. Nearly one third (73.5%) girls had experienced menarche at the age of 10-13 years. During the menarche, 204 (51%) were scared, 73(18.2%) cried, 68 (17%) were embarrassed and only 13.8% were happy. 249 (63.1%) girls experienced pre-menstrual and menstrual problems. 242 (61.4%) girls had missed their schools due to menstruation. Out of them, 54.9% missed a day, 14.4% missed a couple a days and 13.1% missed more than two days in schools per cycle. 74(18.7%) girls did not wash their hand after changing pads.

Table 1 and Table 2 present the frequency and percentage of the knowledge based pre and post test question. Although, there is remarkable change seen in knowledge of pre and post test questions, none had scored 100% correct response in both pre and posttest.

**Table 1.** Knowledge regarding MHM among secondary school girls of Chitwan. (n=400)

**Knowledge regarding MHM among secondary school girls of Chitwan. (n=400)**
| Statements | Pretest correct response |  | Posttest correct response |  |
| --- | --- | --- | --- | --- |
|  | Frequency | (%) | Frequency | (%) |
| Menstruation is the normal and healthy process in female | 210 | 52.5 | 358 | 89.5 |
| Menstruation cycle is related to pregnancy | 376 | 94 | 390 | 97.5 |
| Hormone is the main cause of female menstruation | 323 | 80.8 | 398 | 99.5 |
| Uterus is the main source of bleeding during the menstruation days | 152 | 38 | 343 | 85.8 |
| Normal duration of menstruation flow is 3-5 days | 246 | 61.5 | 339 | 84.8 |
| Normal menstruation cycle for healthy women is 28 days | 210 | 52.5 | 353 | 88.3 |
| 10-15 years is average age of menarche | 372 | 93 | 399 | 99.8 |
| Manage blood flow and maintain hygiene is the main reason to use sanitary pad during menstruation | 393 | 98.3 | 393 | 98.3 |
| In every 4-6 hours sanitary pads should be changed | 311 | 77.8 | 387 | 96.8 |
| Handwashing after pad change is good practice | 394 | 98.5 | 399 | 99.8 |
| Burying and burning is the proper way to dispose a used pad. | 76 | 19 | 205 | 51.3 |

**Table 2.** Knowledge regarding MHM among secondary school girls of Chitwan. (n=400)

| Statements | Pretest correct response |  | Posttest correct response |  |
| --- | --- | --- | --- | --- |
|  | Frequency | (%) | Frequency | (%) |
| Common problems faced before Menarche *<br>Multiple response possible |  |  |  |  |
| Headache | 83 | 20.8 | 239 | 59.8 |
| Constipation/diarrhoea | 25 | 6.3 | 140 | 35 |
| Abdominal bloating | 106 | 26.5 | 208 | 52 |
| Breast tenderness | 181 | 45.3 | 217 | 54.4 |
| Mood swings | 140 | 35 | 198 | 49.5 |
| Common problems faced during menstruation<br>* Multiple response possible |  |  |  |  |
| Cramp, back pain and discomfort | 289 | 72.3 | 359 | 89.8 |
| Loss of appetite , headache and tired | 99 | 24.8 | 258 | 64.5 |
| Full of abdomen and /or difficult to walk | 167 | 41.8 | 233 | 58.3 |
| Remedial measures for problems during menstruation, * Multiple response possible |  |  |  |  |
| Maintain personal Hygiene | 147 | 36.8 | 254 | 63.5 |
| Take rest | 260 | 65 | 308 | 77 |
| Healthy diet | 219 | 54.8 | 270 | 67.5 |
| Light exercise | 110 | 27.5 | 230 | 57.5 |
| Take medicine | 27 | 6.8 | 199 | 49.8 |

In the pretest, 38% of total participants were aware of uterus as the sole source of menstrual bleeding; menstruation is the normal and healthy process in female (52.5%); burying or burning is the correct way to safe disposal of the used pad (19%); Headache (20.8%), constipation or diarrhea(6.3%), bloating (26.5%), Breast tenderness(45.3%) and mood swings(35%) are common problems faced by girls before onset of menstruation; Loss of appetite, headache and tiredness (24.8%), fullness of abdomen, difficulty in walking (41.8%) were common problem faced during menstruation; maintain personal hygiene (36.8%), doing light exercise (27.5%) and taking medicine if necessary (6.8%) are the remedial techniques to deal with the problems during menstruation.

Despite an overall improvement of knowledge in post-test, knowledge on correct way (burying or burning) to dispose pads (51.3%) and knowledge on commonest problems faced during menstruation (constipation/diarrheas(36%), bloating (52%), mood swings (49.5%) and breast tenderness(54.4%)) was not increased as expected.

Table 3 explained about the level of knowledge in pre and posttest. The unsastisfactory level of knowledge among the respondents had decreased from 57.7 % (pretest) to 7.5% (posttest) while there was a significant improvement in satisfactory level of knowledge from 42.3% (pretest) to 92.5% (posttest). This data clearly explained that the satisfactory level of knowledge had improved after menstural education on hygiene management.

**Table 3.** Level of knowledge regarding MHM among secondary school students, Chitwan. (n=400)

| **Study types** | **Level of knowledge** | **Frequency (%)** |
| --- | --- | --- |
| **Pre test** |  |  |
|  | Unsatisfactory | 231(57.7%) |
|  | Satisfactory | 169 (42.3%) |
| **Post test** |  |  |
|  | Unsatisfactory | 30 (7.5%) |
|  | Satisfactory | 370 (92.5%) |

Bivariate logistic regression is used to examine any factors that may be important with knowledge on menstruation hygiene management. Bivariate analysis showed that girls with ethnicity other than Brahmin/Chhetri (OR= 1.7, p=0.014), living in joint family (OR=1.5, p=0.055), studying in public school (OR=2.18, p=0.0001), mother education was primary or secondary level (OR=1.7 p=0.013), and illiterate (OR=2.14 p=0.029), had no or one close female friend (OR=2.5 p=0.002) and had more than one sister (OR=1.65 p=0.033) were possibly more likely to have unsatisfactory level of knowledge on menstruation hygiene management. Unlikely girls’ father whose occupation was business or service (OR=0.52 p=0.008) and girls’ mother whose was business or service (OR=0.55 p=0.045) were possibly less likely to have unsatisfactory level of knowledge on menstruation Hygiene management (Table 4).

**Table 4:** Logistic regression analysis for factors associated with MHM knowledge among secondary school girls of Chitwan (n=400)

| Variables | COR(95% CI) | P-value | AOR(95% CI) | P-value |
| --- | --- | --- | --- | --- |
| **Ethnicity** |  |  |  |  |
| Brahmin/Chhetri | Ref |  |  |  |
| Other than Brahmin/Chhetri | 1.71 | 0.014# | 1.447 | 0.05# |
| **Type of family** |  |  |  |  |
| Nuclear | Ref |  |  |  |
| Joint | 1.54 | 0.055# | 1.604 | 0.048# |
| **Type of school** |  |  |  |  |
| Public | 2.18 | 0.0001# | 1.742 | 0.026# |
| Private | Ref |  |  |  |
| **Father occupation** |  |  |  |  |
| Agriculture | 0.794 | 0.405 | 0.718 | 0.289 |
| Business/service | 0.52 | 0.008# | 0.702 | 0.201 |
| Others(daily wages, ) | **Ref** |  |  |  |
| **Mother education** |  |  |  |  |
| Illiterate | 2.14 | 0.029# | 0.903 | 0.806 |
| Primary/secondary | 1.76 | 0.013# | 1.080 | 0.770 |
| Higher secondary and above | **Ref** |  |  |  |
| **Mother occupation** |  |  |  |  |
| Agriculture | 0.933 | 0.814 | 0.920 | 0.797 |
| Business/service | 0.55 | 0.045# | 0.609 | 0.123 |
| Others | **Ref** |  |  |  |
| **Close female friend** |  |  |  |  |
| 1 or no friend | 2.5 | 0.002# | 2.196 | 0.011# |
| More than 1 | **Ref** |  |  |  |
| **Number of sister** |  |  |  |  |
| No or one sister | **Ref** |  |  |  |
| More than one sister | 1.65 | 0.033# | 1.276 | 0.335 |

(Adjusted with ethnicity status, Types of family, Types of schools, Father Occupation, Mother Occupation, Mother Education, Number of close friend and number of sisters) 1. # denotes statistically significant association, 2. Hosmer and lemeshow test $p=0.583$ , 3. Cox and snell $R.Square=0.092$ , 4. Nagelkerke $R. Square=0.123$ , 4. Ref= Reference category

Multiple logistic regression was used for multivariable analysis to get the final model in this study. Those variables, which were found statistically significant (p<= 0.05) in bivariate analysis, were included into the multivariate regression analysis. The findings revealed that girls whose ethnicity was other than Brahmin/Chhetri (AoR=1.4, p=0.05), lived in joint family (AoR=1.6, p=0.048), studied in public school (AoR=1.7, p=0.026), had one or no close female friend (AoR=2.2, p=0.011) were significantly associated with unsatisfactory level of knowledge as presented in table 4.

Table 5 showed that level of knowledge rank in posttest was statistically significant and higher than level of knowledge rank in pretest (z= 17.129, p=**<**0.001). This result clearly explained that the education on menstruation hygiene to the secondary school girls is one of the effective strategy to enhance level of knowledge on menstruation hygiene.

**Table 5:** Pre and Posttest Knowledge regarding MHM among secondary school student of Chitwan. (n=400)

| Knowledge on MHM | (Q1, Q2, Q3) | Min/Max | Sum rank (-/+) | P-value |
| --- | --- | --- | --- | --- |
| Pre intervention | (10,12,14) | 6/23 | 41 / 76204 | <0.0001## |
| Post intervention | (15,18,20) | 8/24 |  |  |

Most of the participants (90.5%) had experienced cultural restrictions during menstruation. The commonest limitations found during menstruation were visiting temple (97%), performing household puja (95.3%), attending religious functions (88%), cooking in kitchen (64.2%). The least avoided activity was taking food and drinks (18.5%). (Table 6)

**Table 6:** Cultural restriction faced during menstruation. (n=400)

| Variables | Frequency | Percentage (%) |
| --- | --- | --- |
| Ever experienced cultural restriction during menstruation |  |  |
| Yes | 362 | 90.5% |
| No | 38 | 9.5% |
| If yes which ### |  |  |
| Visit temple | 359 | 97% |
| Attend religious function | 352 | 88% |
| Perform household puja | 346 | 95.3% |
| Touch male family member | 127 | 35 % |
| Cook food or enter inside kitchen | 233 | 64.2% |
| Go outside as much as normal | 76 | 20.9% |
| Eat food or drinks of their choice | 67 | 18.5% |
| Sleep in the same bed with others | 85 | 23.4% |
| Sleep in the household members as others | 144 | 39.7% |

*### denotes multiple response.*

## Discussion

This study aimed to assess the level of knowledge on MHM, associated factors and effectiveness of education regarding MHM among the secondary school girls.

The findings of this study revealed that the more than half (57.7%) of secondary school-going girls had unsatisfactory level of knowledge regarding MHM which is comparable with the study in Chitwan, Nepal (60%), Northeast, Ethiopia (49%) and Oyo state, Nigeria (44.1%). ^15,16,17^ The reasons behind the consistency could be possible due to fact that all countries are considered as low and middle income countries hence the constraints faced by girls regarding MHM may be same.

The study done in Dang Nepal (22.3%), Amhare regional state, Ethiopia (9.3%), Sokoto, Nigeria (35%) and South-western Nigeria (30%) found lower proportion of the respondents had unsatisfactory or lower level of knowledge, respectively ^18, 19,20,21^ In contrast to this study, the study done in Doti, Nepal(73.6%), Araihazar, Bangladesh (72.2%), Baghdad, Iraq (74%), Riyan City, Saudi (75.9%) conducted revealed a higher portion of girls had unsatisfactory knowledge about MHM. ^22,23,24, 25^ The difference might be due to change in time and progression of educational supplies as compared to previous time as well as because of divergence scoring system and number of questions related to knowledge assessment for measuring the knowledge level of MHM in different studies.

Current research find out that majority of the girls (85%) were aware about menstruation before menarche which is consistent with the study conducted in Dang, Nepal(89.4%), Northeast, Ethiopia(86.7%) and Southwest, Nigeria (85.4%). ^18, 17, 21^ However, contrasting result with lower percentage was seen in the other studies done in Nagpur, India (36.9%) and South India (39). ^26,27^ This difference might be due to the fact that women from least developed and developing countries might not clearly express their views to educate their girls due to taboo and myth regarding menstruation.

Present study identified that nearly one third (73.5%) girls had experienced menarche at the age of 10-13 years among them 204 (51%) were scared. The result of this study was consistent with the study conducted in Nagpur,India (13 years) and South India (13.4year). ^26, 27^ A study conducted in Amhare regional state, Ethiopia had revealed slightly higher age (14.5 years) of menarche. ^19^ Evidence reveal that menarche is affected by genetic factors, environmental condition, race, body mass index, geographic location, physical activity, health status and psychological factors. ^28^

This study revealed that, majority of the girls (61.4%) missed school days due to menstruation. The similar data was reported by Wash United on MHM day that mentioned 53% of the menstruating Nepali girls are absent in the school during menstruation.^7^ In the pre intervention time, girls had extremely low level of knowledge on “Menstruation as normal and healthy process in female, Uterus as the main source of bleeding during the menstruation days, Normal menstruation cycle of 28 days”. This was supported by a study conducted in Araihazar, Bangladesh ^29^ and Oyo state, Nigeria. ^17^ However studies conducted in Dang Nepal ^18^, Doti, Nepal ^22^, Chitwan Nepal ^30^ found higher proportion of school girls reported correct response on it. The difference might be due to the different study settings in term of urban vs rural, age of the participants selection of private schools vs public schools, school culture and environment regarding menstruation hygiene management.

The present study demonstrates that level of knowledge regarding MHM were relatively low before implementation of the program. After implementation, there was a significant increase in knowledge among the girls (42.3% vs 82.4%). This findings agrees with the results of other studies in Bangladesh (51% Vs 82.2%), ^29^ Korea (42% vs 75%) ^31^, Riyan City Saudi (38% vs 65%), ^25^ Varying increment proportion in above finding may be due time duration: place of educational intervention, difference in number of education, educational level of students, and previous exposure to same program by students.

In this research, there was a difference in the level of knowledge rank in the post test as compared to pretest which was highly statistically significant.(z= 17.13, p=**<0.001**) which explained that the education on menstruation hygiene to the secondary school girls is one of the good strategy to enhance level of knowledge on menstruation hygiene. This results are similar to the results of a study conducted in Southern Haryana, India, ^32^Riyan City Saudi, ^25^ Damietta City, Egypt,^33^ Jumla District, Nepal, ^34^ Pune city, India ^35^

This study identifies that large percentage of respondents experiencing restriction of cultural activities during menstruation. The most common limited activity was to attend and perform religious activities. Similar findings was observed in study conducted in West Bengal, India, ^36^ Chitwan Nepal, ^30^ Kathmandu Valley, Nepal. ^8^ Another study done in rural areas in West Bengal elicited that restricting sour food and not visiting temple have been the most common restriction reported by the participants. ^37^ This is more or less consistent with the present study. Evidence throws light on existing social discriminations, deep–rooted cultural and religious superstitions among girls and women prominently in Hindu religion which force to deteriorating reproductive health. ^8^

In this study, educational status of the mothers was one of the predictors for level of knowledge among participants. This findings agrees with the outcomes of other studies in Dang, Nepal, ^18^Amhare Ethiopia, ^19^ Oyo, state, Nigeria, ^17^ West Bangal, India, ^36^ and Kathmandu, Nepal. ^25^ Girls always feel safer and comfortable while discussing menstruation with their mothers. If mother is illiterate, insufficient knowledge is passed on along with myths and taboos hence mother education showed significant relationship with level of knowledge on MHM. ^27^

Present study highlighted that ethnicity status had role on determining knowledge on MHM. This observation was in agreement with another studies from West Bengal, India, ^36, 37^ Uttar Pradesh, India. ^38^ This could be due to the fact that people from Nepal and India both have a similar context of Hinduism reflecting in caste and ethnicity.

Similarly, this study revealed that, girls who were studying in private schools were more likely to have satisfactory level of knowledge in comparison to the girls studying in public schools which is in line with the study conducted in Dang, Nepal. ^18^ A study conducted in northwestern Nigeria reported contradictory result where type of schools found insignificant regarding knowledge of menstruation. ^39^ The difference might be due to the differences in school policy of different countries settings.

This study also represent that increasing number of close female friends determines the level of knowledge on MHM. This study agrees the findings of the study in Jodhpur India where peer educators or friends were realized as an effective methods of enhancing awareness among adolescents on menstruation hygiene. ^40^ Similarly the study conducted in Damietta City, Egypt revealed that adolescents girls are highly influenced by peer and they are very likely to listen and follow what their peers say. ^33^

The current study revealed that girls who were living in joint family has most likely to have unsatisfactory level of knowledge as compared to the girls living in nuclear family. This could be the fact that the chance of better bonding with mothers and sister, interactions and privacy is high in nuclear family as compared to joint.

This study has revealed several crucial findings and insights regarding MHM for school going girls. Nevertheless, it also has some limitations. First, the posttest findings in this study were taken immediately after health education on MHM provided. Hence, the girls may have reported better knowledge on MHM. Second, information about school absenteeism and cultural restrictions faced were based on self-reporting which may vary accordingly. Although all possible efforts were applied to standardize the educational intervention, it is possible that environmental factors such as differences in the abilities of data collectors and their ability to disseminate study messages may have affected the study. Inspite of such constraints, present study has efforted to explore major findings on MHM which may play crucial role for policy makers in improving reproductive health status in Chitwan, Nepal.

## Conclusion

The majority of the girls had unsatisfactory level of knowledge on MHM. In spite of the fact that large number of girls heard about menstruation before they had menarche, girls were unacceptably unaware about source of bleeding during menstruation, menstruation is a healthy and normal process, common problems faced before and during menstruation and measures to manage these problems effectively. Knowledge on MHM is predisposed by Ethnicity status, Types of family, Number of close female friends, types of schools and mothers’ education. School absenteeism and cultural restriction faced during menstruation is very high. The educational program conducted brings significant change in level of knowledge on different components of MHM. Hence the education intervention demonstrate the suitability and feasibility of implementing a health educational intervention on MHM. Regular and longer educational program may be needed to sustained knowledge and better practice on MHM. Well informed continuous, school educational programme by peer groups should be delivered to the students.

## Data Availability

The dataset used during the study is available from the corresponding author on reasonable request.

## Acknowledgement

The authors would like to express their appreciation to the schools on whose premises the data was gathered. The authors also wish to thank the Education Development and Coordination Unit, Chitwan and Bharatpur Metropolitan for their support during data collection as well as Mr. Shubash Koirala for his support for data analysis. We would also like to thank the study participants and enumerators that took part in the Menstruation Hygiene Management survey.

## Authors’ contribution

GK and NS conceived of the study; participated in the design, coordination, and implementation of all the study field activities; conducted the statistical analysis; and drafted the manuscript; KA conceived of the study, participated in the design, and helped to draft the manuscript. UG, GK and KA participated in the design, conducted the statistical analysis, and helped to draft the manuscript.

## Availability of data and materials

The dataset used during the study is available from the corresponding author on reasonable request.

## Declarations

### Ethics approval and consent to participate

Ethical approval was granted by the Nepal Health Research Council, Institutional Review Board with registration Number 139/2021 P. All participants provided written informed consent before their involvement in the study.

## References

1. Mouse S, Mohan T, Cavill S. Menstruation hygiene matters:A resource for improving menstruation hygiene around the world.Dhaka, Bangladesh.Wateraid;2012. Available from: https://washmatters.wateraid.org/sites/g/files/jkxoof256/files/Menstrual%20hygiene%20matters%20low%20resolution.pdf

2. Amatya P, Ghimire S, Callahan KE, Baral BK, Poudel KC. Practice and lived experience of menstrual exiles (Chhaupadi) among adolescent girls in far-western Nepal. PLoS One. 2018; 13(12):eo208260.

3. Fast facts: nine things you did not know about menstruation [Internet]. Unicef; 2018 May25. Available from: https://www.unicef.org/press-releases/fast-facts-nine-things-you-didnt-know-about-menstruation

4. Reid G, Bruce AW. Urogenital infections in women: can probiotics help?. Postgraduate Medical Journal. 2003; 79:428–432.

5. Dasgupta A, Sarar M. Menstrual hygiene: How hygienic is the adolescent girl? Indian Journal Community Med. 2008; 33(2):77–80.

6. Ten VA. Menstrual Hygiene: A Neglected Condition for the Achievement of Several Millennium Development Goals. European Commission-Europe Aid; 2007.1–24. Available from: https://www.ircwash.org/sites/default/files/Tjon-A-Ten-2007-Menstrual.pdf

7. Public Health Update. Education about menstruation changes everything: Menstruation Hygiene Day [Internet]. Sagunblog; 2020 August 6.Available from: https://publichealthupdate.com/education-about-menstruation-changes-everything-menstrual-hygiene-day-mhday2017-menstruationmatters/

8. Mukharjee A, Lama M, Khakurel U, Jha NA, Ajose F, Acharya S, et al. Perception and pratices of menstruation restrictions among urban adolescent girls and women in Nepal: A cross-sectional survey. BMC Reproductive Health. 2020; 17(81):1–10.

9. Water Aid: Menstrual hygiene and management an issue for adolescent school girls. March 2009. Available from: https://www.wateraid.org/nepal.

10. Morrison J, Basnet M, Bhatta A, Khimbanjar S, Joshi D and Baral S. Menstrual hygiene management in Udayapur and Sindhuli district of Nepal. Nepal. Wateraid; July 2016. 1–52. Available from: https://www.herd.org.np/uploads/frontend/Publications/PublicationsAttachments1/1484557187-Menstrual%20hygiene%20management%20in%20Sindhuli%20and%20Udaypur%20Nepal[1].pdf

11. CBS. National Population and Housing Census, 2011. Social Characterstics Table (Caste/Ethnicity, Mother Tongue and Second Language). Kathamndu: National Planning Commission Secretariates,Central Bureau of Statistics;2014.

12. CBS. Nepal in figures. Kathmandu: National Planning Comission Secretariates, Central Bureau of Statistics; 2012.

13. Education Development and Coordination Unit, Chitwan, 2019

14. Upashe SP, Tekelab T, Mekonnen J. Assessment of knowledge and practice of menstrual hygiene among high school girls on Western Ethiopia. BMC Women’s health. 2015; 15(1):84.

15. Adhikari B, Kandel S. L.Dhungel, and AMandal. Knowledge and Practice regarding menstrual hygiene in rural adolescent girls of Nepal. Kathmandu University Medical Journal. 2007; 5(3): 382–386.

16. Tegegne TK and Sisay MM. Menstruation Hygiene Management and school absenteeism among female adolescent students in Northeast Ethiopia. BMC Public Health. 2014; 14(1):1118.

17. Fehintola FO, Fehintola AO, Aremu AI, Ogunlaja A, Ogunlaja IP. Assessment of knowledge, attitude and practice about menstruation and menstruation hygiene among secondary high school girls in Ogbomoso, Oyo state, Nigeria. International Journal of Reproduction, Contraception, Obstetrics and Gynecology. 2017; 6(5):1726–1732.

18. Bhusal KC, Bhattarai S, Kafle R, Shrestha R, Chhetri P, Adhikari K. Level and Associated Factors of Knowledge regarding Menstrual Hygiene among School-Going Adolescent Girls in Dang District Nepal. Advances in Preventive Medicine. 2020; 2020: 8872119.

19. Gultie DH, Workineh Y. Age of menarche and knowledge about menstrual hygiene management among adolescent school girls in Amhara province, Ethiopia:implecation to health care workers and school teachers. Plos One. 2014; 9(9): e108644.

20. Oche MO, Umar AS, Gana GJ, Ango JT. Menstrual health: the unmet needs of adolescent girls in Sokota, Nigeria. Scientific Research and Essays. 2012; 7(3):410–418.

21. Aluko OO, Oluya OM, Olaleye OA, Olajuyin AA, Olabintan TF, Oloruntoba-Oju OI. Knowledge and Menstrual hygiene practices among adolescent in senior secondary schools in Ile Ife, south-western, Nigeria. Journal of Water, Sanitation and Hygiene for Development. 2014; 4(2).248–256.

22. Yadav NR, Joshi S, Poudel R, Pandeya P. Knowledge, attitude, and practice on menstrual hygiene management among school adolescents. Journal of Nepal Health Research Council. 2018; 15(3):212–216.

23. Haque ES, Rahman M, Itsuko Mutaharu M, Sakisaka K. The effect of a school-based educational intervention on menstrual health: an intervention study among adolescent girls in Bangladesh. BMJ Open. 2014; 4(7): e004607.

24. Sadiq AM, Salih AA. Knowledge and practice of adolescent females about menstruation in Baghdad. Journal of General Practice. 2013; 2(1): 1–4.

25. AI Ayafi AS. Factors associated with health behavior regarding menstruation among Saudi intermediate school girls in Riyadh City (Doctoral Dissertation, Master thesis). Nursing College, King Saud University. 1999. Available on: https://scholar.google.com/scholar?q=Ayafi+SA++Factors+associated+with+health+behavior+regarding+menstruation+among+Saudi+intermediate+school+girls+in+Riyadh+city++[+Master+thesis+].++Nursing+College,+King+Saud+University+,++1999+

26. Thakre BS, Thakre SS, Reddy M, Rathi N, Pathak K, Ughade S. Menstrual hygiene:knowledge and practice among adolescent school girls of saoner, Nagpur district. Journal of Clinical and Diagnostic Research. 2011; 5(5):1027–1033.

27. Omidyar S, Begum K. Factors influencing hygienic practices during menses among girls from south India-a cross sectional study. International Journal of Collaborative Research on Internal Medicine and Public Health. 2010; 2(12):411–423.

28. Tehrni RF, Mirmiran P, Gholami R, Nazanin M, Azizi F. Factors influencing Menarcheal age: results from the cohort of Tehran lipid and glucose study. International Journal of Endocrinology and Metabolism. June 2014; 12(3):e16130.

29. Haque SE, Rahman M, Itsuko K, Mutahara M, Sakisaka K. The effect of a school-based educational intervention on menstrual health:An intervention study among adolescent girls in Bangladesh. BMJ Open. 2014; 4(7):e004607.

30. Neupane MS, Sharma K, Bista AP, Subedi S, Lamichhane S. Knowledge on menstruation and menstrual hygiene practices among adolescent girls of selected schools of Chitwan. Journal of Chitwn Medical College. 2020;10(1):69–73.

31. Min J, Ahn S. Effects of menstrual self-management education program on knowledge and behavior of menstrual self-management in high school girls. Korean Journal women Health Nursing. 2018; 24:310–321.

32. Singh A, Gupta V, Agrawal D, Goyal P, Singh M, Lukhmana S. A cross-sectional study to investigate the impact of focused group discussion on menstrual hygiene among rural school girls of Southern Haryana, India. Journal of Education and Health Promotion. Oct 2020;9: 260.

33. Mohamad HA,El-Karmalawy EM, Gida NI, Hafez FE. Effects of health education program on menstrual practices among secondary school girls. Port Said Scientific Journal of Nursing. June 2018; 5(1): 27–52.

34. Dahal A. Acharya KP. Effectiveness of information education and communication on menstrual hygiene among adolescent school girls of Jumla District. Journal of Nobel Medical College. 2019; 8:4–9.

35. Prema S, Dhandapani D, Prakash D, Gawade S. effectiveness of planned health teaching on knowledge and self –reported practices of menstrual hygiene among visually impaired adolescent girls in selected blind schools of Pune city. International Journal of Advances in Nursing Management. 2020;8:53–56.

36. Sarkar I, Dobe M. Dasgupta A, Basu R, Shahbabu B. Determinants of menstrual hygiene among school going adolescent girls in a rural area of West Bengal. Journal of Family Medicine and Primary Care. 2017;6(3):583–588.

37. Ray S, Dasgupta A. Determinants of Menstrual hygiene among adolescent girls:A multivariate analysis. National Journal of Community Medicine. 2012; 3(02):294–301.

38. Malhotra A, Goli S, Coates S, Mosquera-Vasquez M. Factors associated with knowledge, attitudes, and hygiene practices during menstruation among adolescent girls in Utter Pradesh. Waterlines. July 2016; 35(3):277–305.

39. Lawan UM, Yusuf NB, Musa AB. Menstruation and menstruation hygiene amongst adolescent school girls in Kano, Northwestern Nigeria. African Journal of Reproductive Health. 2010; 14(3):201–207.

40. Dwivedi R, Sharm C, Bhardwaj P, Singh K, Joshi N, Shrama PP. Effect of peer educator-PRAGATI(Peer Action for Group Awareness Through Intervention) on Knowledge, attitude, and practice of menstrual hygiene in adolescent school girls. Journal of Family Medicine and Primary Care. July 2020; 9(7):3593–3599.

